# SARS-CoV-2 Prevalence and Seroprevalence among Healthcare Workers in Belgian Hospitals: Baseline Results of a Prospective Cohort Study

**DOI:** 10.1101/2020.10.03.20204545

**Authors:** Laure Mortgat, Cyril Barbezange, Natalie Fischer, Leo Heyndrickx, Veronik Hutse, Isabelle Thomas, Bea Vuylsteke, Kevin Arien, Isabelle Desombere, Els Duysburgh

**Affiliations:** Department of Epidemiology and Public Health, Sciensano, Brussels, Belgium; European Programme for Intervention Epidemiology Training (EPIET), European Centre for Disease Prevention and Control (ECDC), Stockholm, Sweden; Department of Infectious diseases in humans, Sciensano, Brussels, Belgium; European Public Health Microbiology Training (EUPHEM), European Centre for Disease Prevention and Control (ECDC), Stockholm, Sweden; Department of Biomedical Sciences, Institute of Tropical Medicine, Antwerp, Belgium; Department of Public Health, Institute of Tropical Medicine, Antwerp, Belgium; University of Antwerp

## Abstract

**Background:** Given the current SARS-CoV-2 pandemic and the occurrence of a second wave, assessing the burden of disease among health care workers (HCWs) is crucial. We aim to document the prevalence of SARS-CoV-2 and the seroprevalence of anti-SARS-CoV-2 IgG among HCWs in Belgian hospitals, and to study potential risk factors for the infection in order to guide infection prevention and control (IPC) measures in healthcare institutions.

**Methods:** We performed a cross-sectional analysis of the baseline results (April 22 - April 26) of an ongoing cohort study. All staff who were present in the hospital during the sampling period and whose profession involved contact with patients were eligible. Fourteen hospitals across Belgium and 50 HCW per hospital were randomly selected. RT-qPCR was performed to detect SARS-CoV-2 RNA on nasopharyngeal swabs, and a semi-quantitative IgG ELISA was used to detect anti-SARS-CoV-2 antibodies in sera. Individual characteristics likely to be associated with seropositivity were collected using an online questionnaire.

**Findings:** 698 participants completed the questionnaire; 80.8% were women, median age was 39.5, and 58.5% were nurses. Samples were collected on all 699 participants. The weighted anti-SARS-CoV-2 IgG seroprevalence was 7.7% (95%CI, 4.7%-12.2%), while 1.1% (95%CI, 0.4%-3.0%) of PCR results were positive. Unprotected contact with a confirmed case was the only factor associated with seropositivity (PR 2.16, 95% CI, 1.4-3.2).

**Interpretation:** Most Belgian HCW did not show evidence of SARS-CoV-2 infection by late April 2020, and unprotected contact was the most important risk factor. This confirms the importance of widespread availability of protective equipment and use of adequate IPC measures in hospital settings.

## Introduction

Early December 2019, a novel coronavirus, the severe acute respiratory syndrome coronavirus 2 (SARS-CoV-2) emerged in Wuhan, China, causing the condition called Coronavirus disease 2019 (COVID-19) with symptoms ranging from mild respiratory symptoms to a serious, sometimes life-threatening pneumonia. The virus rapidly spread to more than 200 countries and territories worldwide, and as of 14^th^ of September 2020, resulted in more than 28 million cases including almost 917 500 deaths.^1^

Health care workers (HCW) play a critical role in the clinical management of patients and in ensuring that adequate infection prevention and control (IPC) measures are implemented in health care facilities. As such, HCW not only represent a highly exposed population, but also play a crucial role in the chain of transmission, as they are in close contact with vulnerable patients at high risk for COVID-19.

Belgium was hit hard during the first wave of the epidemic. The first imported case was reported on February 3^rd^, and local transmission was identified early March.^2^ Until the 14^th^ of September 2020, around 93 500 confirmed cases and 19 500 hospital admissions were reported. Half of the 10 000 deaths due to COVID-19 occurred in the hospital.^3^ As in many other countries, testing strategies have changed over the months, partly in response to available resources. Initially, possible cases* in the general population were only tested if they required hospitalisation. Similarly, HCW were only tested for SARS-CoV-2 if they had respiratory symptoms accompanied by fever; by mid-April, the presence of fever was no longer a prerequisite for testing. Many pauci-symptomatic or asymptomatic cases were therefore never identified. Since May 4^th^, all possible cases** can be tested. In parallel, since mid-March, containment measures such as social distancing, closure of non-essential facilities and borders, or restrictions on gatherings have been progressively put in place by the National Security Council in the attempt to limit the spread of the virus and reduce the pressure on the healthcare system.

Measuring the SARS-CoV-2 infection among HCW is important not only to estimate the current burden of disease in this population, but also to reduce secondary virus transmission within health care settings. At this stage of the epidemic and in the context of ongoing transmission, it is crucial to properly assess the proportion of HCW who is still immunologically naïve, and who was infected and developed potentially protective immunity. To date, seroprevalence studies were only carried-out in single hospitals in Belgium, and a nation-wide overview is lacking.

We initiated a prospective cohort study to investigate and follow up SARS-CoV-2 prevalence and seroprevalence among hospital HCW in Belgium, in order to support and guide IPC measures in hospitals and healthcare resources planning. We additionally aim to identify potential risk factors for the infection and to generate insights into the clinical presentation of the disease by assessing reported symptoms and the proportion of asymptomatic infections. In this paper we describe the baseline findings of this ongoing study.

## Methods

### Study design, participants and sampling methods (ClinicalTrials.gov Identifier: NCT04373889)

This prospective cohort study includes seven time points, with a follow-up every two weeks during the first month, then monthly for four months until the end of September 2020. In this paper, we report on the baseline data collected between the 22^nd^ and the 26^th^ of April 2020 among a representative sample of 699 HCW in Belgium.

Recruitment of hospitals and participants started in early April. All medical and paramedical staff who were present in the hospital during the sampling period and whose profession involved contact with patients were eligible. Temporary staff was excluded. In each hospital, a local coordinator was designated to ensure appropriate communication with the participants and the researchers and to support the logistics of the study.

At the time of the study design, anti-SARS-CoV-2 IgG seroprevalence data were scarce. To calculate the sample size needed, we therefore opted for a conservative approach and used an estimated seroprevalence of 50%. Considering a precision of 5%, a sample of 385 participants was required. We performed a two-stage cluster sampling assuming a design effect of two to account for the loss of precision due to correlation within the clusters. Thus we required 48.15 subjects per cluster, which we rounded upwards to 50, reaching a total sample size of 800 HCW. In a first stage, we selected a random sample of 16 out of the 104 Belgian general hospitals, with a probability of sampling proportional to size. The number of beds was taken as a proxy for size and number of HCW. In a second stage, we randomly selected 50 HCW in each hospital, using the staffing records as sampling frame. If the selected HCW did not accept to participate, the next HCW on the list was contacted.

After having given informed consent, study participants were asked to provide a nasopharyngeal swab and a blood sample to test for the presence of SARS-CoV-2 RNA and anti-SARS-CoV-2 IgG antibodies respectively. Samples were collected by trained staff, as recommended by Sciensano. In addition, participants were invited to complete a questionnaire using an online tool (LimeSurvey). The questionnaire could be self-completed, in the hospital or at home, or completed with the help of the local study coordinator. Samples and questionnaires were collected over a period of five days.

The baseline questionnaire collected information on basic socio-demographic characteristics (date of birth and sex), health information (comorbidities, past or current COVID-19 compatible symptoms, previous COVID-19 diagnosis, use of angiotensin-converting-enzyme (ACE) inhibitors), and work related information (occupation, years of experience, work schedule, working in a COVID-19 dedicated unit, contact with a confirmed case with and without using recommended precautions). Participants who failed to complete the questionnaire received up to two reminders from the local study coordinator.

Each participant was assigned a unique study code by the local coordinator, who is the only person able to link patient name and study code. Laboratory samples and questionnaires were pseudonymised and linked through this code. This code was also used by the researchers team to communicate test results to the participants in a confidential way, using a dedicated phone line.

### Laboratory methods

Biological samples were transported to the services Viral Diseases and Immune Response of Sciensano, the Belgian institute of public health, and to the Virology Laboratory of the Institute of Tropical Medicine, Antwerp (ITM). Real-time reverse transcription polymerase chain reaction (RT-qPCR) targeting the E gene was performed to detect SARS-CoV-2 RNA on nasopharyngeal swabs according to the method described by Corman et al.,^4^ using a Ct cut-off of 40. Presence of IgG antibodies in sera was tested using a commercially available semi-quantitative test from Euroimmun (anti-SARS-CoV-2 IgG ELISA, reference EI 2606-9601 G, Medizinische Labordiagnostika AG). This test reveals the presence of IgG directed to the S1 domain of the SARS-CoV-2 spike protein. Based on reported and *in house* validations, it has a specificity of 98.6% and a sensitivity at 14 days post clinical illness onset of 95.9%.^5–7^ Results are comparable with the performance characteristics reported by the manufacturer.^8^ Sera were considered positive at an S/N ratio ≥1.1, as suggested by the manufacturer.

### Data analysis

Because it is our intention to infer the findings of the study sample to the population of all Belgian HCW, we corrected the analyses for the two-stage sampling design, by applying inverse probability weighting and survey statistics functions. For descriptive data analysis, including estimations of prevalence and seroprevalence, we calculated proportions with a 95% confidence interval. We used Poisson regression adjusted for the sampling probability to identify association between anti-SARS-CoV-2 IgG seropositivity and health and work related risk factors as well as symptoms. Associations were first assessed in univariate models. Factors and symptoms significant at the 0.10 level were included in the final model, along with age and sex as potential confounders. The variable “having had a previous COVID-19 diagnostic” was excluded from the model, as infection is a pre-requisite for antibody production and because of the high correlation with symptoms. A separate model was constructed to estimate the predictive value of symptoms. A backward stepwise model selection was performed, comparing the Akaike Information Criterion (AIC) of each model, including possible interaction, to determine the goodness of fit. Data were analysed using STATA/SE 16.1.

### Ethics considerations

Written informed consent was obtained from all HCW before enrolment in the study. To guarantee confidentiality, study laboratory results and questionnaires were pseudonymised using unique study codes. The study was approved by the Medical Ethics Committee of the University Hospital Ghent (reference: B6702020000036).

### Role of the funding source

This study was funded by Sciensano, the Belgian institute of public health, Brussels, Belgium. Sciensano was involved in all stages of the study, from conception and implementation to analysis and reporting.

## Results

### General characteristics

The baseline sampling took place between April 22^nd^ and April 26^th^ and involved 699 participants in 14 hospitals geographically spread across Belgium. Due to logistical and time constraints, the other two hospitals and one participant joined the study at subsequent timepoints, and are therefore not included in this baseline analysis. One participant did not complete the questionnaire.

The participants were predominantly women, had a median age of 39.5, and more than half of them were nurses (58.5%). Participants characteristics are presented in table 1.

**Table 1:** General characteristics of the study participants (N=698)

| **Healthcare workers characteristics** |  |
| --- | --- |
| Age, median (IQR, range) (N=698) | 39.5 (32-49, 20-67) |
| Sex (N=698) |  |
| • Male, n (%) | 134 (19.2) |
| • Female, n (%) | 564 (80.8) |
| Profession (N=696) |  |
| • Medical doctor, n (%) | 155 (22.3) |
| • Nurse, n (%) | 407 (58.5) |
| • Other paramedical staff <sup>a</sup>, n (%) | 134 (19.2) |
| Years of experience, median (IQR, range) (N=688) | 14 (7-23, 0-45) |
| Working in a COVID ward, n (%) (N=682) | 334 (49.0) |

COVID-19: Coronavirus Disease 2019, IQR: interquartile range, N: total number, n: number in that category
<sup>a</sup> Include among others physiotherapists, care auxiliaries, occupational therapists, technicians, psychologists, dieticians, and midwives.

### Prevalence and seroprevalence of SARS-CoV-2

Crude sample proportions and weighted estimates are presented in table 2. We discuss here weighted results only. Out of the 699 participants tested, 8 had a positive PCR resulting in a SARS-CoV-2 prevalence of 1.1% (95% CI 0.4%-3.0%). Four of those testing positive had experienced symptoms with an onset less than 14 days ago, one was never symptomatic, and three had symptoms dating back more than three weeks. Twenty-seven participants (3.5%, 95% CI 1.5%-8.0%) reported a previous positive PCR test result. Among them, four (15.5%) did not show a detectable IgG response. For one of them, diagnosis was obtained only two days earlier, while for the three other participants, diagnosis occurred more than three weeks earlier.

**Table 2:**
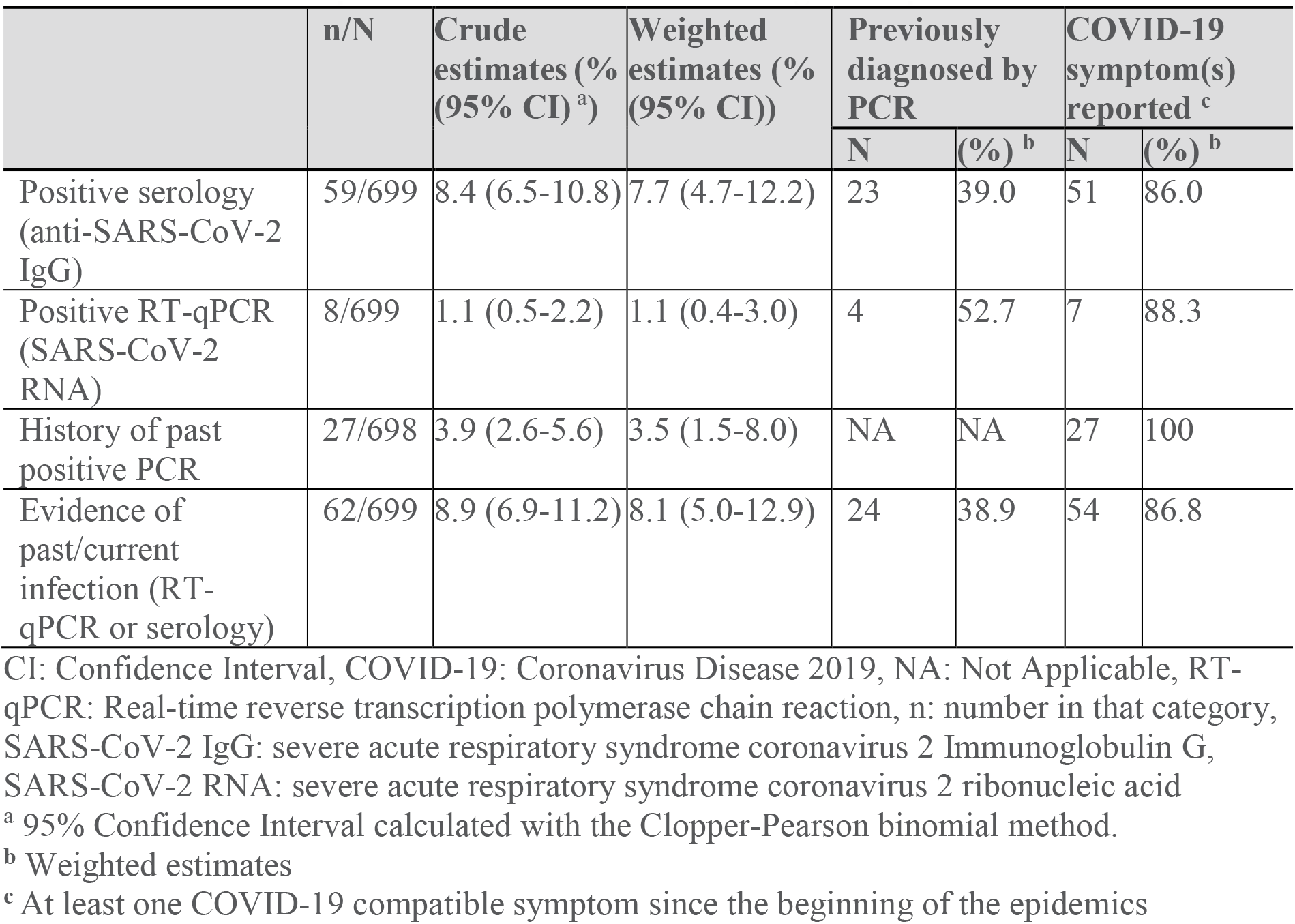
SARS-CoV-2 prevalence, anti-SARS-CoV-2 IgG seroprevalence.

|  | n/N | Crude estimates (%) (95% CI)<sup>a</sup> | Weighted estimates (%) (95% CI) | Previously diagnosed by PCR |  | COVID-19 symptom(s) reported<sup>c</sup> |  |
| --- | --- | --- | --- | --- | --- | --- | --- |
|  |  |  |  | N | (%)<sup>b</sup> | N | (%)<sup>b</sup> |
| Positive serology (anti-SARS-CoV-2 IgG) | 59/699 | 8.4 (6.5-10.8) | 7.7 (4.7-12.2) | 23 | 39.0 | 51 | 86.0 |
| Positive RT-qPCR (SARS-CoV-2 RNA) | 8/699 | 1.1 (0.5-2.2) | 1.1 (0.4-3.0) | 4 | 52.7 | 7 | 88.3 |
| History of past positive PCR | 27/698 | 3.9 (2.6-5.6) | 3.5 (1.5-8.0) | NA | NA | 27 | 100 |
| Evidence of past/current infection (RT-qPCR or serology) | 62/699 | 8.9 (6.9-11.2) | 8.1 (5.0-12.9) | 24 | 38.9 | 54 | 86.8 |

CI: Confidence Interval, COVID-19: Coronavirus Disease 2019, NA: Not Applicable, RT-qPCR: Real-time reverse transcription polymerase chain reaction, n: number in that category, SARS-CoV-2 IgG: severe acute respiratory syndrome coronavirus 2 Immunoglobulin G, SARS-CoV-2 RNA: severe acute respiratory syndrome coronavirus 2 ribonucleic acid
<sup>a</sup> 95% Confidence Interval calculated with the Clopper-Pearson binomial method.
<sup>b</sup> Weighted estimates
<sup>c</sup> At least one COVID-19 compatible symptom since the beginning of the epidemics

Anti-SARS-CoV-2 IgG antibodies were found in 59 samples, resulting in a seroprevalence of 7.7% (95% CI 4.7%-12.2%). Among the seropositive individuals, 36 (61.0%) never had a positive test before, among which 8 (22.9%) did not report any COVID-19 compatible symptoms since the beginning of the epidemic. In seropositive participants who reported previous COVID-19 compatible symptoms, median time since onset of symptoms was 27 days, with a range between 11 and 56 days, except for one participant who mentioned symptoms dating back two days prior to sampling. In total, 62 participants (8.1%, 95% CI 5.0%-12.9%) showed evidence of active or previous infection (PCR+ and/or serology+).

### Demographic, health related and work related characteristics associated with SARS-CoV-2 seropositivity

Based on univariate analysis (table 3), age and sex were not significantly different between HCW with and without anti-SARS-CoV-2 IgG antibodies. Similarly, none of the comorbidities tested (cardiovascular disease, hypertension, diabetes, chronic lung disease, immunodeficiency etc.), were significantly associated with SARS-CoV-2 seropositivity, neither was smoking nor the intake of ACE inhibitors and Sartans (data not shown). Among work-related risk factors, an unprotected contact with a confirmed COVID-19 patient was the only factor associated with seropositivity with a risk ratio of 2.16 (95% CI 1.44-3.23). In multivariable analysis, when adjusting for age and sex, the risk ratio remained similar at 2.11 (95% CI 1.36-3.28).

**Table 3:** Demographic and work related factors associated with anti-SARS-CoV-2 IgG seropositivity, univariate analysis.

|  | Positive serology (n=59) |  | Negative serology (n=640) |  | Weighted prevalence ratio (95% CI) |
| --- | --- | --- | --- | --- | --- |
| Demographic characteristics | Number | % | Number | % |  |
| Sex (N=698) |  |  |  |  |  |
| • female | 42 | 71.2 | 522 | 87.7 | ref. |
| • male | 17 | 28.8 | 117 | 18.3 | 1.55 (0.71-3.38) |
| Age group (N=698) |  |  |  |  |  |
| • 20-34 years | 26 | 44.1 | 216 | 33.8 | ref. |
| • 35-49 years | 22 | 37.3 | 285 | 44.6 | 0.70 (0.34-1.46) |
| • 50 years and above | 11 | 18.6 | 138 | 21.6 | 0.67 (0.27-1.69) |
| **Work related factors** |  |  |  |  |  |
| Profession (N=696) |  |  |  |  |  |
| • Other paramedic | 11 | 18.6 | 123 | 19.3 | ref. |
| • Nurse | 35 | 59.3 | 372 | 58.4 | 0.97 (0.51-1.81) |
| • Medical doctor | 13 | 22.0 | 142 | 22.3 | 0.95 (0.33-2.68) |
| ≤5 years of experience (N=688) | 18 | 30.5 | 123 | 19.6 | 1.59 (0.64-3.94) |
| Schedule (N=693) |  |  |  |  |  |
| • Only night shifts | 1 | 1.7 | 21 | 3.3 | ref. |
| • Only day shifts | 33 | 56.9 | 348 | 54.8 | 2.75 (0.35-21.31) |
| • Mixed: day and night shifts | 24 | 41.4 | 266 | 41.9 | 2.75 (0.30-25.34) |
| Full-time (N=613) | 34 | 67.4 | 332 | 58.9 | 1.30 (0.65-2.62) |
| COVID ward (N=698) |  |  |  |  |  |
| • Working in COVID ICU unit | 8 | 13.6 | 94 | 14.7 | 0.85 (0.35-2.09) |
| • Working in COVID non-ICU unit | 20 | 33.9 | 248 | 38.8 | 0.85 (0.46-1.59) |
| Contact with a confirmed COVID-19 patient |  |  |  |  |  |
| • With recommended precautions (N=627) | 38 | 73.1 | 412 | 71.7 | 1.07 (0.61-1.87) |
| • Without recommended precautions (N=575) | 24 | 50 | 161 | 30.6 | 2.16 (1.44-3.23) |

CI: Confidence Interval, COVID-19: Coronavirus Disease 2019, ICU: Intensive Care Unit, N: total number, n: number in that category

### Symptoms as predictors of SARS-CoV-2 seropositivity

Most of the assessed symptoms were significantly associated with anti-SARS-CoV-2 IgG seropositivity (table 4) in univariate analysis. Among them, anosmia or ageusia, pain, fever and fatigue showed the highest relative risk. All symptoms associated with seropositivity with a p value<0.10 were retained in the final multivariable model, except for skin rash that was removed to achieve the best fit (lowest AIC).When adjusted for other symptoms and for age and sex, only anosmia or ageusia remained statistically significant with a prevalence ratio of 6.71 (95% CI 3.19-14.12).

**Table 4:** Symptoms as predictors of SARS-CoV-2 seropositivity.

|  | SARS-CoV-2 IgG seroprevalence |  |  |  | Weighted prevalence ratio (95% CI) |
| --- | --- | --- | --- | --- | --- |
|  | Symptom present |  | Symptom absent |  |  |
| Symptoms (N=698) | n/N | % | n/N | % |  |
| At least one symptom | 51/421 | 12.1 | 8/277 | 2.9 | 4.04 (2.06-7.92) |
| • Loss of smell and/or taste | 31/51 | 60.8 | 28/647 | 4.3 | 14.88 (8.07-27.44) |
| • Pain | 25/72 | 34.8 | 34/626 | 5.4 | 7.10 (4.08-12.36) |
| • Fever/chills | 27/86 | 31.4 | 32/612 | 5.2 | 6.49 (4.63-9.10) |
| • Fatigue/weakness | 33/125 | 26.4 | 26/573 | 4.5 | 5.59 (3.50-8.91) |
| • Dyspnea | 21/77 | 27.3 | 38/621 | 6.1 | 3.95 (2.36-6.62) |
| • Cough | 33/106 | 19.9 | 26/532 | 4.9 | 3.75 (2.07-6.78) |
| • Diarrhea and/or vomiting | 18/88 | 21.6 | 40/610 | 6.6 | 3.53 (2.52-4.94) |
| • Headache | 35/222 | 15.8 | 24/476 | 5.0 | 3.51 (1.97-6.26) |
| • Skin rash | 5/21 | 23.8 | 54/677 | 8.0 | 3.02 (1.31-7.00) |
| • Running nose | 23/168 | 13.7 | 36/530 | 6.8 | 1.82 (0.99-3.37) |
| • Sore throat | 19/185 | 10.3 | 40/513 | 7.8 | 1.24 (0.72-2.14) |
| • Mental confusion/irritability | 1/8 | 12.5 | 58/690 | 8.4 | 1.35 (0.19-9.43) |

CI: Confidence Interval, N: total number, n: number in that category, CI: Confidence Interval, SARS-CoV-2 IgG: severe acute respiratory syndrome coronavirus 2 Immunoglobulin G

## Discussion

Based on our findings, an estimated 7.7% of Belgian hospital HCW had developed IgG antibodies against SARS-CoV-2 by late April 2020. Including those with a positive RT-qPCR, it can be estimated that 8.1% had been in contact with the virus by then. To the best of our knowledge, the present study is the first report on seroprevalence in HCW at national level, using a representative sample of HCW. Our results are in-line with the 6.4% seroprevalence reported in another Belgian study, where 3056 staff of one tertiary hospital were screened for anti-SARS-CoV-2 antibodies during almost the same period.^9^ In other seroprevalence studies conducted in hospital HCW in Europe, findings were very heterogeneous and varied between countries, regions, and sampling period. In Germany, the estimated seroprevalence among HCW was lower and varied between 1% and 2.7% depending on the region.^10–12^ On the contrary, in highly affected regions as Madrid and Lombardy, seroprevalence reached up to 31.6%^13^ and 43%^14^ respectively, while in other parts of both countries, seroprevalence was either similar^15^ or lower than ours.^16,17^ A higher seroprevalence was also reported in Sweden (19.1%),^18^ where the policy response to COVID-19 was not as stringent as in other European countries. It should be noted that all of these studies, unlike ours, were conducted either in a single centre, or in a sample of healthy volunteers.

As it can be assumed that HCW are potentially highly exposed, we expected seroprevalence in this population to be higher than among the general population. At the end of April, seroprevalence in the Belgian population was estimated at 6.0% in a study using residual sera,^19^ and at 4.7% in a study among blood donors.^20^ In the first study, samples originating from hospitals were excluded, while in the second, the ‘healthy volunteer effect’ (i.e. blood donors) most probably led to an underestimation. Our findings reveal that, at the time of the study, Belgian HCW were more affected by SARS-CoV-2 than the general population, but not to a great extent.

The only identified risk factor for seroconversion was the contact with a confirmed case without using recommended precautions. This result further stresses the importance of strict IPC measures and use of PPE as long as the virus is circulating in order to limit hospital-associated infections. Working in a COVID-19 unit was not identified as a risk factor for seropositivity, probably because greater risk awareness leads to more careful precautions. Olfactory and gustatory symptoms were the strongest predictors for seropositivity. This finding is consistent with other studies that show a strong association with anti-SARS-CoV-2 IgG seropositivity, in HCW^9,14,15,18,21^ and among the general population,^22^ as well as with acute SARS-CoV-2 infection.^13,23,24^ In Belgium, these symptoms have been included as “major symptoms” in the definition of a possible case since the 4th of May.^25^

Our study has several limitations. First, the characteristics of the ELISA test used might have led to a few diagnostic flaws. Follow-up of the HCWs in this study together with additional laboratory testing on stored samples (plaque reduction neutralisation) will likely give further insights in the extent of such diagnostic accuracy issues. Second, we cannot rule out the possibility of selection bias, as some of the initially selected participants may have refused to participate or were on sick leave at the time of the first sampling. The latter could have underestimated seroprevalence. The number of refusals was however reported to be very low, as at time of recruitment demand for testing in HCW was very high. Also, two of the selected hospitals entered the cohort in May and were not included in these baseline results, reducing the statistical power of the study. Recent evidence^26,27^ suggest that antibodies tend to wane in the early convalescent phase (8 weeks after infection), which means that HCW infected in the beginning of the epidemic might not have been detected. This could explain why in four participants who reported a previous positive PCR, no antibodies were detected. Finally, it should be noted that serological studies do not provide a complete picture of the immune response to SARS-CoV-2, in which T-cell mediated immunity seems to play an important role, especially in the long term.^28,29^

Still, the main strength of this study lies in the fact that its design allows generalisation of its results to all Belgian HCW. Additionally, since current knowledge on the protective potential of antibodies is still lacking, the longitudinal part of the study will enable monitoring the persistence of SARS-CoV-2 antibodies and detecting new infections, seroconversions and possibly reinfections.

We have shown that the majority of HCW in Belgium showed no evidence of previous contact with the virus in late April. This indicates that herd immunity, certainly if defined by the presence of antibodies, should not be relied upon to reduce virus transmission, but that widespread availability of PPE and use of adequate IPC measures should be reinforced and maintained.

## Data Availability

Due to confidentiality agreements, supporting data cannot be made openly available.

## Authors contribution

Mortgat and Duysburgh had full access to all the data in the study and accept responsibility for the decision to submit for publication.

Concept, design, protocol writing: Mortgat, Duysburgh, Arien Logistical coordination: Mortgat, Duysburgh

Administrative, technical, or material support: all authors

Biological samples collection, transport and analysis: Arien, Barbezange, Desombere, Fisher, Heyndrickx, Hutse, Thomas

Epidemiological data collection, cleaning and analysis: Mortgat, Duysburgh Drafting of manuscript: Mortgat

Manuscript revision: Duysburgh, Barbezange, Vuylsteke, Arien, Fischer, Hutse, Thomas, Desombere

Funding: Duysburgh, Desombere Statistical analysis: Mortgat, Duysburgh Supervision: Duysburgh

## Declaration of interest

None

## Ethic committee approval

The study was approved by the Medical Ethics Committee of the University Hospital Ghent on the 8^th^ of April 2020 (reference: B6702020000036).

## Acknowledgements

We want to thank each of the participating hospitals and their local coordination team for their logistical support:

AZ Jan Yperman Ziekenhuis: Study nurse Melissa Devos and hospital hygiene team led by Dirk Vanrenterghem
Prof Dr Erika Vlieghe (Antwerp University Hospital / Institute of Tropical Medicine) – Dr Patrick Soentjens (Institute of Tropical Medicine / Antwerp University Hospital)
AZ Damiaan ziekenhuis, Oostende
AZ Sint-Lucas Brugge: Dr. Johan Robbrecht, Head of Department of Clinical Biology, Lislot Mommerency, Coordinator of Clinical Studies, Larissa Staelens, Study Coordinator
AZ West: Leen Pollet, MD, Klaas Vandevyvere, MD, Lieve Debruyne, hospital hygiene nurse
CHIREC-hôpital de Waterloo-Braine-l’Alleud
Cliniques de l’Europe, Bruxelles: Matthieu Pierre Rutgers, MD, Sarah Debray, MD, Olivier Lang (nurse), Isabelle Schmidt (nurse), Sandrine Van Wilderode (nurse) et Sara Yagirian (nurse)
Clinique Saint Pierre Ottignies: Dr Grégoire Wieërs, Dr Valérie Selosse, Catherine Georges, Isabelle Vanbellinghen, Martine Gérard
Ghent University Hospital, Dept Internal Medicine: Steven Callens, MD/PhD, Sophie Van Herreweghe
Jessa Ziekenhuis : Apr Biol Sara Vijgen and Dr Koen Magerman, in collaboration with MENSURA : Dr. Nele Van Loon and Mathieu Verbrugghe
Mariaziekenhuis Noord-Limburg
Vivalia – Centre Hospitalier de l’Ardenne: Gerald Malempré, Sylvie Thomas, Marie André, MD, Pierre Yves Machurot, MD.

For her help in providing results feedback to participants, we also thank Veerle Boonen, MS, Department of Epidemiology and Public Health, Sciensano, Brussels, Belgium.

For their technical assistance: Department of Infectious diseases in humans, Sciensano, Brussels, Belgium:

Caroline Rodeghiero, Fabienne Jurion, Assia Hamouda, Ilham Fdillate, Reinout Van Eycken, Mona Abady, Aurélie Francart, Jeannine Weyckmans, Vera Verburgh, Sophie Lamoral.

For their logistical assistance in preparing the kits: Kim Borighem, Vincent Marteau, Pascale Elsocht; and in sample management: Bart De Logi, Nathalie Stassen.

Thanks to Sooria Balasegaram, MD; for her support, review and input at all stages of the study. Our gratitude finally goes to all the study participants.

## Footnotes

* As of the 11^th^ of March, possible cases were defined as a person with symptoms of acute infection of the lower or upper respiratory tract that appear or worsen when the patient has chronic respiratory symptoms.

** As of May 4^th^ and until today, a possible case is defined as a person with at least one of the major symptoms (cough, dyspnoea, chest pain, anosmia or dysgeusia) with acute onset and without other obvious cause; or at least two of the minor symptoms (fever, pain, muscle aches; fatigue; rhinitis; sore throat; headache; anorexia; watery diarrhoea, acute confusion, sudden fall) with no other obvious causes; or an aggravation of chronic respiratory symptoms (COPD, asthma, chronic cough…), without another obvious cause.

## Notes

### Competing Interest Statement

The authors have declared no competing interest.

### Clinical Trial

ClinicalTrials.gov Identifier: NCT04373889

### Author Declarations

Ethical approval was granted by the Medical Ethics Committee of the University Hospital Ghent on the 8th of April 2020 (reference: B6702020000036).

